# Clinical Decision Support Opportunities among Clinicians Caring for People Receiving Invasive Mechanical Ventilation: A Cross-sectional Survey

**DOI:** 10.1101/2024.05.20.24307549

**Authors:** Gary E. Weissman, Nicholas S. Bishop, Benjamin E. Schmid, Vanessa Madden, Romain Pirracchio, Jacob C. Jentzer, Kathryn A. Riman, Dustin C. Krutsinger, Bruno DiGiovine, Lyle H. Ungar, Scott D. Halpern, Catherine L. Auriemma, Thomas S. Valley, Meeta Prasad Kerlin

**Affiliations:** Palliative and Advanced Illness Research (PAIR) Center, University of Pennsylvania Perelman School of Medicine, Philadelphia, PA, USA; Department of Anesthesia and Perioperative Care, University of California at San Francisco, San Francisco, CA, USA; Department of Cardiovascular Medicine, The Mayo Clinic, Rochester, MN, USA; Department of Critical Care Medicine, University of Pittsburgh, Pittsburgh, PA, USA; Pulmonary, Critical Care & Sleep Medicine Division, Department of Medicine, University of Nebraska, Omaha, NE, USA; Division of Pulmonary, Critical Care, and Sleep Medicine, Trinity Health, Livonia, MI, USA; Department of Computer and Information Science, University of Pennsylvania, Philadelphia, PA, USA; Pulmonary and Critical Care Medicine, Department of Medicine, University of Michigan, Ann Arbor, MI, USA

## Abstract

Treatment decisions for patients receiving invasive mechanical ventilation (IMV) are complex and depend simultaneously on the current ventilator settings, the function of multiple interrelated organ systems, and other treatments. An artificial intelligence (AI)-based clinical decision support system (CDSS) offers a promising approach to alleviate uncertainty in this complex management and provide personalized treatment recommendations. However, little is known about clinician preferences for which treatment decisions clinicians think would most benefit from a CDSS. Therefore, we conducted a cross-sectional electronic survey of practicing physicians, nurses, advanced practice providers, and respiratory therapists to identify key treatment decisions in IMV care. We sent the survey instrument to 132 clinicians across six geographically diverse health systems. Among 51 respondents (39% response rate), there were 24 (47%) physicians, 7 (14%) registered nurses, 8 (16%) advanced practice providers, and 12 (24%) respiratory therapists. Participants were from five US states including Pennsylvania, California, Michigan, Minnesota, and Nebraska. At least 50% of participants identified 18 distinct treatment decisions for IMV care as *very important* or *absolutely essential*, including many that were outside of the ventilator settings themselves. The highest agreement about importance was for the decision to extubate (N=51, 100%), and the decisions to conduct a spontaneous awakening trial (N=48, 94%) and spontaneous breathing trial (N=48, 94%). The highest agreement about a decision being *not important* or only *slightly important* was for the shape of the inspiratory flow pattern (N=15, 29%). These findings underscore the scope and complexity of clinical decision making across multidisciplinary teams in caring for people receiving IMV. Furthermore, they underscore the gap between important clinician-identified decisions and existing IMV CDSSs that provide suggestions for at most 5 ventilator settings without support for other related decisions. Future work is needed to identify which of these decisions might be most appropriate for inclusion in a CDSS to support IMV management.

## 1 Introduction

People with respiratory failure requiring invasive mechanical ventilation (IMV) are at high risk for morbidity and mortality, and incur large health care costs (Bellani et al., 2016; Stefan et al., 2013; Teno et al., 2022; Wunsch et al., 2010). Caring for people receiving IMV is highly complex and requires making over a dozen interrelated treatment decisions that can change in a matter of minutes (Figure 1). These decisions include the ventilator settings themselves, such as mode, set tidal volume, set respiratory rate, and fraction of inspired oxygen (FiO2), among others. But these decisions are closely linked with other medical interventions, the function of other organ systems, and the underlying cause of respiratory failure. For example, infusions of analgesic, sedative, vasopressor, and neuromuscular blockading medications are essential elements of caring for people receiving IMV and their administration and dosing are tightly interrelated with choices about optimal ventilator settings. Other important categories of treatments include the use of inhaled pulmonary vasodilators, prone positioning, and the decision to extubate, among others. When dealing with an individual patient, there may be multiple sets of treatment decisions that could achieve the same goal. But existing treatment guidelines focus on a single or handful of these decisions and do not offer precise recommendations for all how to make optimal care decisions while accounting for all of these elements simultaneously.

**Figure 1.**
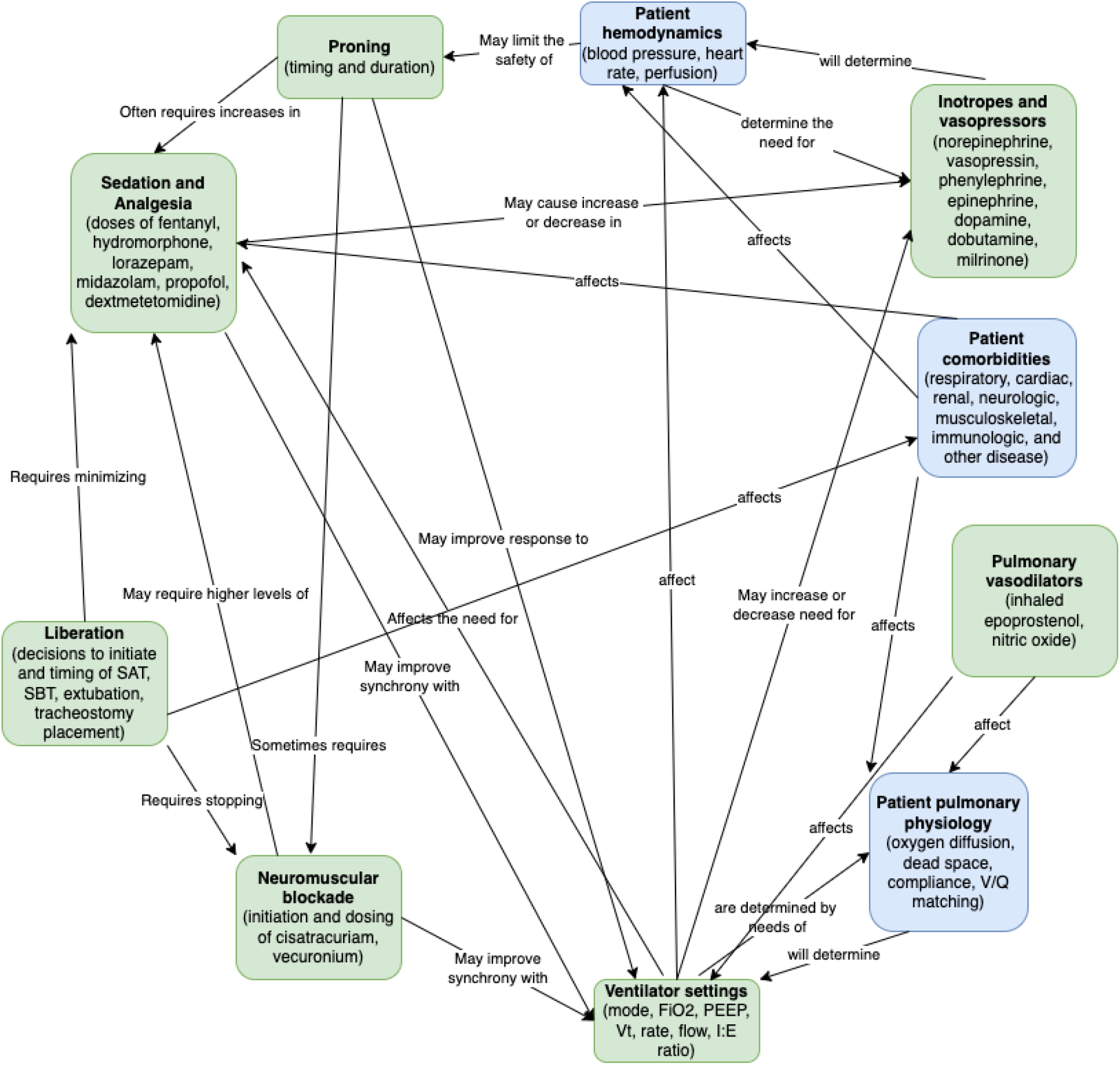
A subset of the many complex, interrelated, and simultaneous categories of information that clinicians face related to treatment decisions (green) and patient physiology (blue) when caring for people receiving invasive mechanical ventilation.

Because of this complexity and uncertainty, there is significant variation in care delivery and outcomes for people receiving IMV with large inequities between rural and urban hospitals (Balas et al., 2022; Harlan et al., 2024; Kalhan et al., 2006; Mikkelsen et al., 2008; Qadir et al., 2021). By reducing unwarranted variation in care patterns, an artificial intelligence (AI) clinical decision support system (CDSS) could ameliorate uncertainty in these complex decisions (Chen et al., 2022; Gandhi et al., 2023) support personalized treatment recommendations,(Shah et al., 2021; Yildirim et al., 2024a), and overcome known barriers to optimal IMV care (Dexter and Schleyer, 2022; Fitzpatrick and Ellingsen, 2013; Yildirim et al., 2024b). Previously developed IMV CDSSs have focused on only one or a handful treatment decisions, and none provides suggestions for more than five ventilator settings simultaneously (Tehrani and Roum, 2008). Therefore, we sought to elicit clinicians’ assessments of the importance of key IMV treatment decisions. We conducted a cross-sectional electronic survey of physicians, nurses, advanced practice providers, and respiratory therapists across diverse practice settings to identify the most important IMV-related decisions and inform the development of future CDSSs.

## 2 Methods

### 2.1 Instrument Development

We developed an electronic survey instrument with three components (see *Appendix*). First, we asked participants to report their demographic information, including information about their clinical role, prior experience with clinical decision support for IMV, and location of practice. Second, we asked participants to rate the importance of 20 different treatment decisions related to the management of patients with IMV. The list of decisions was determined based on expert clinician guidance and revised by the study team for clarity and consistency. This list was presented in two chunks of 10 each to facilitate understanding by each participant. The first bundle included the decisions most likely to be relevant for all patients receiving IMV and the second bundle included those only relevant for some patients receiving IMV. The order of the specific decisions was randomized within each bundle to avoid ordering effects. Third, participants were asked to indicate their willingness to participate in a future Delphi study about IMV CDSS. The instrument was prepared using the web-based Qualtrics (Provo, Utah) survey platform.

### 2.2 Population and Recruitment

Investigators across six geographically diverse health systems (Penn Medicine, University of Nebraska, University of Michigan, Trinity Health, University of California San Francisco, Mayo Clinic Rochester) compiled a list of potential survey participants and their email addresses from each institution. These lists included physicians, nurses, advanced practice providers (nurse practitioners and physician assistants), and respiratory therapists. An initial email request for participation was sent to the entire aggregated list and followed by two weekly reminders sent to non-respondents between April and May 2024.

### 2.3 Analysis

We reported counts and proportions of the responses across each decision and rating. The study was approved by the Institutional Review Board of the University of Pennsylvania. All analyses were conducted using the R Language for Statistical Computing (R Core Team, Vienna, Austria) version 4.2.

## 3 Results

Of 132 candidate participants who were sent emails, 51 (39%) completed the survey (Figure 2). Among these 51 respondents, most were physicians (N=24, 47%), worked in a medical ICU (N=40, 78%), and practiced in Pennsylvania (N=28, 55%). Complete respondent characteristics are reported in Table 1.

**Table 1:** Characteristics of study participants. Abbreviations: ICU = intensive care unit, LTACH = Long-term acute care hospital, CRNP = Certified registered nurse practitioner, PA = Physician assistant.

| Participant Characteristic | N (%) |
| --- | --- |
| **Total participants** | 51 (100%) |
| **Gender** |  |
| Female | 22 (43.1%) |
| Male | 29 (56.9%) |
| **Race** |  |
| Asian | 1 (2.0%) |
| Black or African American | 1 (2.0%) |
| More than one race | 1 (2.0%) |
| Not Reported | 1 (2.0%) |
| White | 47 (92.2%) |
| **Ethnicity** |  |
| No, not of Hispanic, Latino/a/x, or Spanish origin | 49 (96.1%) |
| Not Reported | 1 (2.0%) |
| Yes, of Hispanic, Latino/a/x, or Spanish origin | 1 (2.0%) |
| **State of Practice** |  |
| CA (California) | 5 (9.8%) |
| MI (Michigan) | 11 (21.6%) |
| MN (Minnesota) | 4 (7.8%) |
| NE (Nebraska) | 3 (5.9%) |
| PA (Pennsylvania) | 28 (54.9%) |
| **Primary Clinical Role** |  |
| Advanced Practice Provider (CRNP or PA) | 8 (15.7%) |
| Physician (MD or DO) | 24 (47.1%) |
| Registered Nurse | 7 (13.7%) |
| Respiratory Therapist | 12 (23.5%) |
| **Primary Practice Site** |  |
| Medical ICU | 40 (78.4%) |
| General ICU | 12 (23.5%) |
| Cardiac ICU | 10 (19.6%) |
| Surgical ICU | 10 (19.6%) |
| Neurologic ICU | 6 (11.8%) |
| Emergency Department | 3 (5.9%) |
| Operating Room | 3 (5.9%) |
| LTACH | 1 (2.0%) |

**Figure 2.**
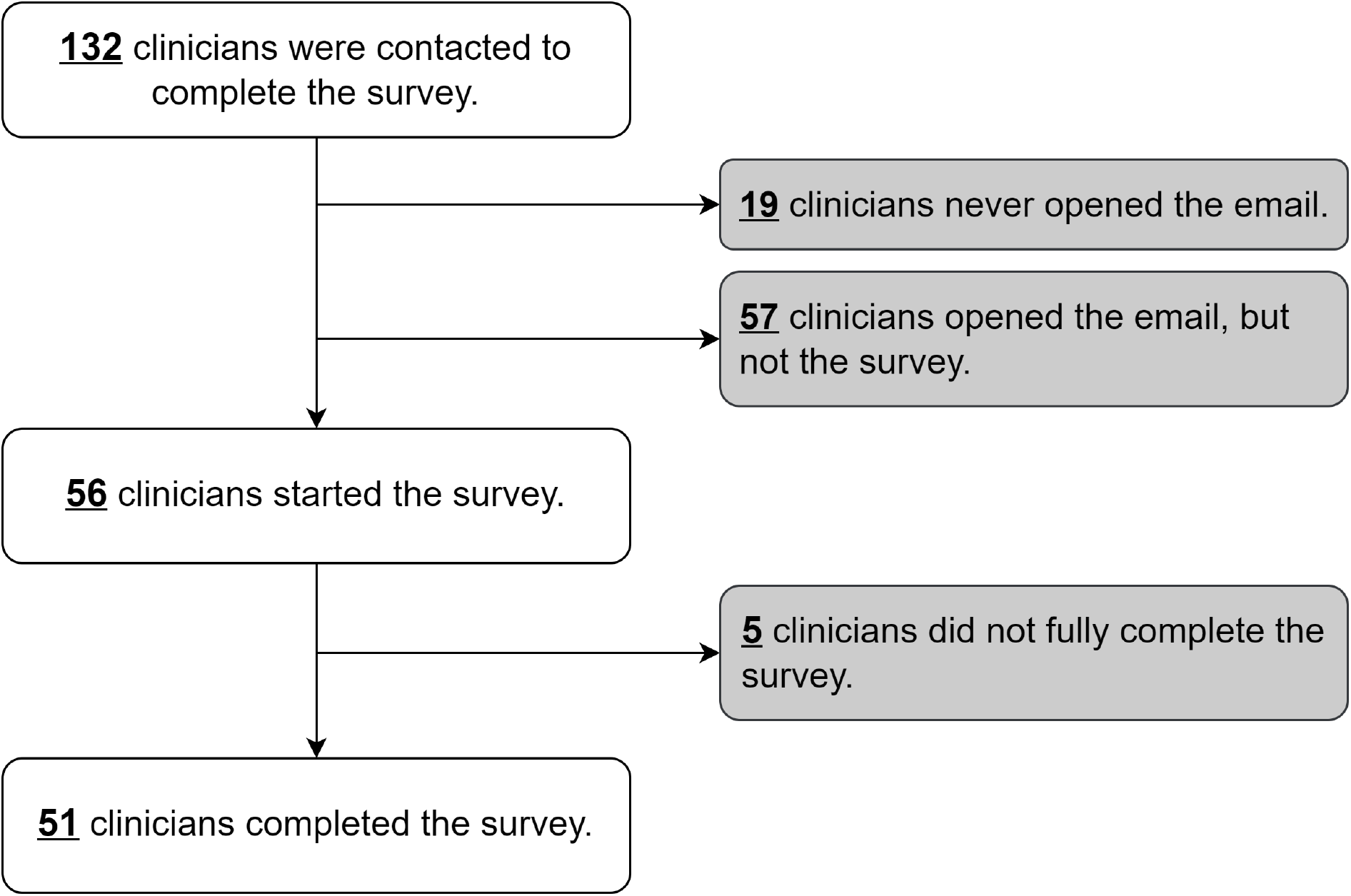
Recruitment of clinicians and participation in the study.

Among the 20 candidate treatment decisions, the decision to extubate was most consistently rated *very important* or *absolutely essential* (N=51, 100%). The highest agreement about a decision being *not important* or *slightly important* was for the shape of the inspiratory flow pattern (N=15, 29%).

At a consensus threshold of 50%, participants identified 18 decisions as *very important* or *absolutely essential* (Figure 3). Among these were decisions about extubation, sedation, analgesia, prone positioning, tracheostomy placement, and vasopressors, in addition to various settings on the ventilator itself. At a threshold of 70%, participants identified 14 such decisions.

**Figure 3.**
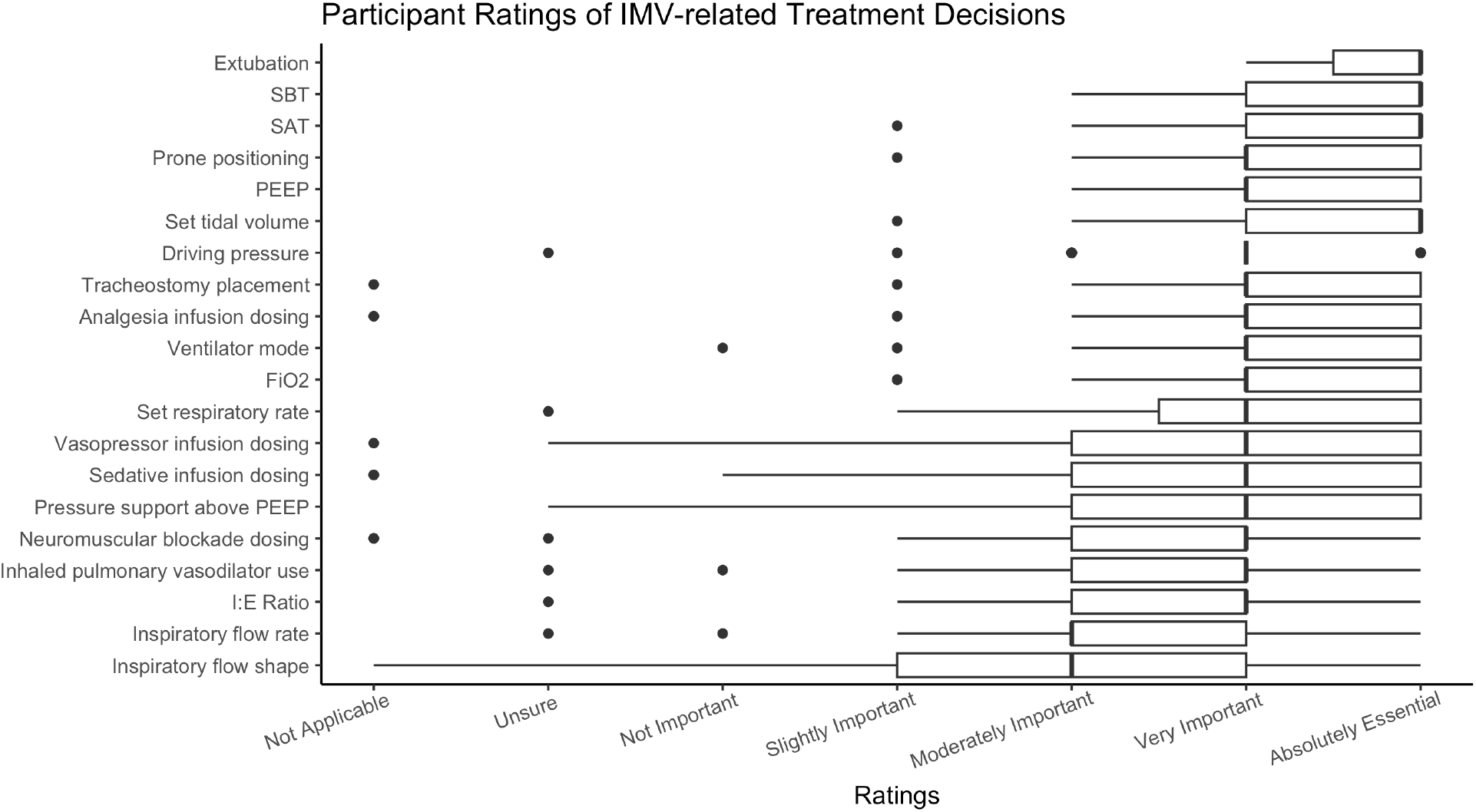
Box and whisker plots showing the distribution of ratings for each treatment decision. Decisions are ordered along the y-axis in descending order based on the proportion of respondents who indicated that decision was *very important* or *absolutely essential*. Abbreviations: IMV = invasive mechanical ventilation, SBT = spontaneous breathing trial, SAT = spontaneous awakening trial, PEEP = positive end expiratory pressure, FiO2 = fraction of inspired oxygen, I:E = inspiratory to expiratory time.

19 (37%) of respondents reported having used a CDSS related to IMV management in the past. 48 (94%) expressed interest in using a CDSS for IMV management if it were shown to be safe, effective, and equitable. And 39 (76%) expressed interest in participating in a future Delphi study related to IMV CDSSs.

## 4 Discussion

We conducted an electronic survey of practicing clinicians who care for people receiving IMV to assess their ratings of the importance of associated treatment decisions. This is the first such assessment and provides essential data for the future development of CDSSs for IMV management in a diverse and multidisciplinary environment. Notably, at a consensus threshold of 50%, we found that clinicians identified 18 important decisions, many of which were outside of the ventilator settings themselves. These findings highlight the limitations of currently available IMV CDSSs that provide suggestions for at most 5 ventilator settings without support for other related and important decisions.

Clinician recognition of an IMV-related related treatment decision as important is a necessary step in the development of future CDSSs. However, it is not sufficient to determine if a decision should be included in a future CDSS. For example, inclusion of all 18 decisions identified as important here could lead to information overload for an end-user at the bedside. Thus, future work is needed to rank the relative importance of these and other decisions and to quantify the tradeoffs between their inclusion and potential CDSS overcrowding. Clinicians may also prefer that some decisions be better addressed through non-CDSS means, such as through pop-up alerts, nudges, checklists, or other strategies designed to inform care practices.

This study has important limitations. First, participants were restricted to rating only the 20 decisions included in the initial survey instrument. Other related decisions such as early mobilization, ventilator dyssynchrony identification, and others, were not included in order to limit the burden on participants. Therefore, the importance of many decisions not presented in this instrument would not have been captured in this study. Future studies should evaluate the importance of these and other treatment decisions. Second, we did not include pharmacists, physical therapists, occupational therapists, or participants from other professions whose opinions are important to IMV management. Third, the survey may suffer from non-response bias in that clinicians more knowledgeable or more opinionated about this topic may have been more likely to respond. Clinicians with less knowledge may be especially likely to benefit from an IMV CDSS but their ratings of the most important decisions would not have been captured in this design.

In conclusion, clinicians practicing in geographically and clinically diverse settings, and from across professions, identify a large number of important decisions relevant for the care of people with IMV. These important decisions extend outside of just the ventilator settings and exceed the decision support capacity of currently available systems. Future work on the development of CDSSs for IMV management should incorporate the preferences of end-users to ensure clinical relevance at the bedside.

## Data Availability

All data produced in the present study are available upon reasonable request to the authors.

## 5. Acknowledgements

We are grateful to all of the busy clinicians who took time to complete this survey and share their opinions and expertise.

## A Appendix Questions included in the electronic survey instrument

1. Electronic consent
2. Please indicate your primary clinical discipline
  - Physician (MD or DO)
  - Registered Nurse
  - Advanced Practice Provider (CRNP or PA)
  - Respiratory Therapist
  - Pharmacist
  - Other
3. In which type of setting do you usually care for people receiving invasive mechanical ventilation (check all that apply)
  - Intensive care unit, general
  - Medical intensive care unit
  - Surgical intensive care unit
  - Neurologic intensive care unit
  - Cardiac intensive care unit
  - Operating room/procedure suite
  - Emergency department
  - Other
4. In which state do you primarily see patients
5. Which best describe(s) your gender identity?
  - No, not of Hispanic, Latino/a/x, or Spanish origin
  - Yes, Mexican, Mexican American, Chicano/a/x
  - Yes, Peurto Rican
  - Yes, Cuban
  - Yes, Another Hispanic, Latino/a/x or Spanish origin
6. Are you Hispanic, Latino/a/x, or of Spanish origin?
7. What is your race? (select all that apply)
  - White
  - Black or African American
  - American Indian or Alaskan Native
  - Asian
  - Native Hawaiian or Pacific Islander
  - Prefer to specify/Other
8. Have you interacted with a clinical decision support tool related to ventilator management before in your practice?
  - Yes
  - No
9. Would you be interested in using a clinical decision support tool related to ventilator management in your practice if it were shown to be effective, safe, and equitable?
  - Yes
  - No
10. Imagine you are working in your usual setting caring for patients who receive invasive mechanical ventilation. The following treatment decisions are typically made for most patients who receive invasive mechanical ventilation. Select the choice that best represents how important, on average, that decision is in caring for your patients?
  - Ventilator mode (e.g. assist control/volume cycled, pressure support)
  - Set tidal volume (mL)
  - Set respiratory rate (breaths per minute)
  - Pressure support above PEEP (cm H2O)
  - Positive end espiratory pressure (PEEP)
  - Fraction of inspired oxygen (FiO2)
  - Spontaneous breathing trial (SBT) initiation
  - Spontaneous awakening trial (e.g. sedation interruption, SAT) initiation
  - Decision to extubate
  - Inspiratory flow rate (Liters per minute)
11. Imagine you are working in your usual setting caring for patients who receive invasive mechanical ventilation. The following treatment decisions are made for certain patients who receive invasive mechanical ventilation. When the decision is relevant for a patient, select the choice that best represents how important, on average, that decision is in caring for your patients?
  - Inhaled pulmonary vasodilators (e.g. epoprostenol or nitric oxide)
  - Decision to place in prone position
  - Decision to place a tracheostomy
  - Vasopressor infusion dosing (e.g. norepinephrine or epinephrine)
  - Analgesia infusion dosing (e.g. fentanyl, morphine, or hydromorphone)
  - Sedative infusion dosing (e.g. propofol or lorazepam)
  - Neuromuscular blockade dosing (e.g. cisatracurium)
  - Driving pressure (plateau pressure - PEEP)
  - Inspiratory flow shape (e.g. ramp or decelerating)
  - Inspiratory to expiratory time (I:E) ratio
12. Please click below if you are willing to be contacted about participating in a future Delphi panel (a virtual and anonymous process primarily conducted through electronic surveys to elicit your feedback and preferences) related to clinical decision support and invasive mechanical ventilation. You would receive a cash incentive for participating in this future panel.

